# Temporal Associations of Plasma Levels of the Secreted Phospholipase A_2_ Family and Mortality in Severe COVID-19

**DOI:** 10.1101/2022.11.21.22282595

**Authors:** Eric Lu, Aki Hara, Shudong Sun, Brian Hallmark, Justin M. Snider, Michael C. Seeds, Joseph C. Watkins, Charles E. McCall, Hao Helen Zhang, Guang Yao, Floyd H. Chilton

## Abstract

Previous research suggests that group IIA secreted phospholipase A_2_(sPLA_2_-IIA) plays a role in and predicts severe COVID-19 disease. The current study reanalyzed a longitudinal proteomic data set to determine the temporal (days 0, 3 and 7) relationship between the levels of several members of a family of sPLA_2_ isoforms and the severity of COVID-19 in 214 ICU patients. The levels of six secreted PLA_2_ isoforms, sPLA_2_-IIA, sPLA_2_-V, sPLA_2_-X, sPLA_2_-IB, sPLA_2_-IIC, and sPLA_2_-XVI, increased over the first 7 ICU days in those who succumbed to the disease. sPLA_2_-IIA outperformed top ranked cytokines and chemokines as predictors of patient outcome. A decision tree corroborated these results with day 0 to day 3 kinetic changes of sPLA_2_-IIA that separated the death and severe categories from the mild category and increases from day 3 to day 7 significantly enriched the lethal category. In contrast, there was a time-dependent decrease in sPLA_2_-IID and sPLA_2_-XIIB in patients with severe or lethal disease, and these two isoforms were at higher levels in mild patients. Taken together, proteomic analysis revealed temporal sPLA_2_ patterns that reflect the critical roles of sPLA_2_ isoforms in severe COVID-19 disease.

## 1 Introduction

Disseminated extrapulmonary COVID-19 disease leads to organ and tissue dysfunction and high mortality rates (1-5). Major gaps persist in understanding the molecular biology driving COVID-19 diffuse vascular and organ inflammation. Metabolomic/lipidomic studies of circulating lipids align disseminated COVID-19 disease syndrome severity with reduced phospholipids and elevated levels of lyso-phospholipids (lyso-PL) and unesterified unsaturated fatty acids (UFA) (6, 7). This metabolic profile suggested the hydrolysis of circulating or cellular phospholipids by one or more phospholipase A_2_ (PLA_2_) activities was associated with COVID-19 disease pathobiology.

The secreted PLA_2_ (sPLA_2_) family consists of low molecular weight proteins (14–15 kDa) that require millimolar Ca^2+^ for their catalytic activity and primarily hydrolyze target phospholipids in the extracellular space or on the outer layers of cellular membranes (8, 9). The mammalian sPLA_2_ family presently contains 10 known catalytically active isoforms (**Figure 1A**; sPLA_2_-IB, sPLA_2_-IIA, sPLA_2_-IIC, sPLA2-II-D, sPLA_2_-II-E, sPLA_2_-II-F, sPLA_2_-III, sPLA_2_-V, sPLA_2_-X, sPLA_2_-XIIA) and one catalytically inactive isoform (sPLA_2_-XIIB) with distinct tissue/cellular distributions, phospholipid substrate specificities, and diverse biology.

**Figure 1:**
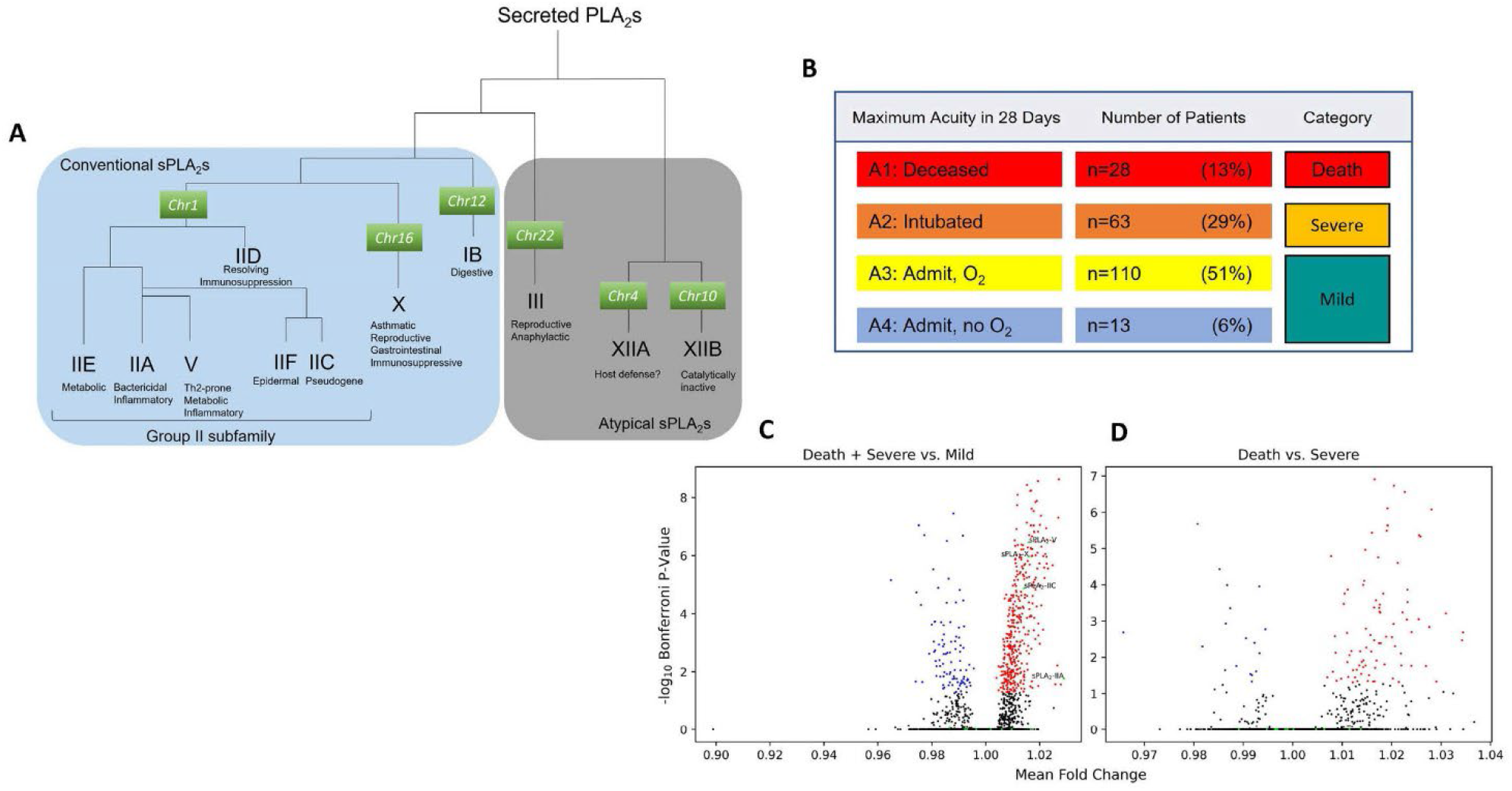
Secreted PLA_2_ Family, Distribution of Clinical Acuity Categories of COVID-19 Cohort and Volcano Plots Showing Baseline Fold Change Comparisons between Acuity Groups. **A**. A phylogenetic tree of sPLA_2_ family and representative functions of each isoform. Conventional sPLA_2_s (group I/II/V/X) are closely related low-molecular-weight enzymes with a highly conserved Ca^2+^-binding loop and a His/Asp catalytic dyad as well as conserved disulfide bonds, while atypical sPLA_2_s (group III and XII) are each classified into distinct classes. **B**. Distribution of the COVID-19-infected cohort with, day 0, day 3, and possibly day 7 timepoints used in downstream proteomic analyses. Each patient was observed for their most severe acuity within 28 days post-enrollment. Categories referenced in later analyses are defined as Death, Severe, and Mild for A1, A2, and (A3-A4), respectively. **C**. Volcano plot displaying p-values of Bonferroni-adjusted, two-sided independent t-tests and average fold-changes between the baseline (day 0) protein levels of combined Death + Severe vs. Mild-category patients, and **D** Death vs. Severe-category patients. All sPLA_2_ isoforms are colored green and those with statistically significant differences between groups are labeled by name.

Experimental and clinical evidence in the 1980s revealed that sPLA_2_ plays a critical role in multiple organ failure lethality. More specifically, Vadas recognized a mechanistic relationship between high circulating sPLA_2_ activity and hypotension and respiratory distress syndrome during gram-negative bacterial septic shock (10). Thereafter identified as sPLA_2_-IIA, temporal measurements of sPLA_2_-IIA in the ICU emerged as an early and accurate predictor of multiple organ failure syndrome severity and lethality in patient subgroups (11-13). Specifically, patients who succumbed to lethal organ failure from sepsis had persistently increased circulating sPLA_2_-IIA activity whereas survivors experienced a tapering of levels.

We discovered that sPLA_2_-IIA may function in and predict severe COVID-19 disease outcomes (14). Using three machine learning algorithms, sPLA_2_-IIA was identified as the key variable among 80 clinical indices with the capacity to stratify severe COVID-19 survivors from deceased victims.

Absence of temporal kinetics of sPLA_2_-IIA levels limited the previous study, as did lack of sPLA_2_ isoform selective ELISA kits to detect and quantify other sPLA_2_ isoforms. The current study addresses both limitations using the clinical dataset of Filbin et al (15) and plasma proteomics analysis of eight sPLA_2_ isoforms and an intracellular calcium-dependent PLA_2_ (16, 17) collected on days 0, 3, and 7 of patients entering the ICU. Temporal correlations of several sPLA_2_ isoforms with COVID-19 deaths recapitulated earlier sepsis studies. Furthermore, differences in the kinetics of sPLA_2_ isoforms known to be proinflammatory versus immunosuppressive also emphasized the potential divergent roles sPLA_2_ family members in COVID-19 severity.

## 2 Methods

### 2.1 Dataset

Proteomic and clinical index datasets generated by Filbin et al. were obtained from Mendeley Data (15). The datasets included 384 patients enrolled from the Emergency Department (ED) of the Massachusetts General Hospital (MGH) with 306 patients positive for SARS-CoV-2. Patient acuity levels were classified (A1-A5) based on the World Health Organization (WHO) Ordinal Outcomes Scale and determined by their severest condition within 28 days post-enrollment. Among the 306 COVID patients, 214 had blood samples obtained at two time points (days 0 and 3) and 128 at three timepoints (days 0, 3, and 7). Plasma levels of 5,201 proteins were measured using the SomaScan platform and expressed as Relative Fluorescence Units (RFUs). Our reanalysis included the 214 COVID patients with at least two blood samples, of which 28 patients died (A1), 63 required intubations (A2), 110 required supplemental oxygen (A3), and 13 were without supplemental oxygen during hospitalization (A4). Patients discharged from ED without hospitalization and blood samples at day 3 (A5) were not included in our reanalysis. A1, A2, and (A3-A4) were grouped into the categories of death (n=28), severe (n=63), and mild (n=123), respectively. Protein level RFUs were log_2_ transformed. ∆1 and ∆2 values were calculated as the differences between log_2_ protein levels on days 3 and 0 and days 7 and 3, respectively.

### 2.2 Statistics

Significance of protein level differences between sample groups were calculated using independent two-sided t-tests with Bonferroni correction to correct for multiple hypothesis testing. Adjusted p-values were binned into 0-0.0001, 0.0001-0.001, 0.001-0.01, 0.01-0.05, and 0.05-1 and labeled in plots with four to zero asterisks, respectively.

### 2.3 Decision Tree Analysis

A classification model used the ∆1 and ∆2 values of sPLA_2_-IIA through recursive partitioning (decision tree) to classify patient categories (death, severe, mild) using the R package rpart (18). The minimum number of samples at a node to allow a split was set to 20 and the complexity assessed by the lowest 10-fold cross-validation error. Patients with missing ∆2 values were not propagated through decisions involving ∆2 (use surrogate=0). Splits calculations used Gini impurity.

Performance assessed using ROCs and AUCs were calculated using the pROC (19) package.

## 3 Results

**Figure 1B** shows the four clinical primary acuity levels of 214 COVID-19 patients with day 0, day 3, and possibly day 7 timepoints used in downstream proteomic analyses. **Figure 1C and Supplementary Table 1** compares fold changes and statistical significance of measured proteins in deceased patients + intubated patients versus milder disease. Among 492 statistically significant elevated proteins, sPLA_2_-IIA, sPLA_2_-V, sPLA_2_-X and sPLA_2-_IIC ranked 8^th^, 227^th^, 231^st^, and 337^th^, respectively. **Figure 1D** shows that, when assessing only baseline levels, none of these sPLA_2_ isoforms significantly differed between patients who died compared to severe or intubated.

Figure 2. compares levels of the nine measurable PLA_2_ isoforms including sPLA_2_-IB, sPLA_2_-IIA, sPLA_2_-IIC, sPLA_2_-IID, sPLA_2_-IIE, sPLA_2_-V, sPLA_2_-X, sPLA_2_-XIIB, and PLA_2_-XVI. Six of the nine isoforms, sPLA_2_-IIA, sPLA_2_-V, sPLA_2_-X, sPLA_2_-IB, sPLA_2_-IIC, and PLA_2_-XVI, increased over time in non survivors. In contrast, ventilated survivors begin to resolve elevations between day 3 and day 7. Mild patients had reduced levels of sPLA_2_-IIA, sPLA_2_-IIC, sPLA_2_-IIE, sPLA_2_-V, sPLA_2_-X, and PLA_2_-XVI over time. In contrast, there was a time-dependent decrease in sPLA_2_-IID and sPLA_2_-XIIB in patients who died and in severe patients who lived. These two isoforms also were found at higher levels in less severe patients.

**Figure 3A** is a spaghetti plot of the longitudinal flow of sPLA_2_-IIA and confirms PLA_2_-IIA as a key protein associated with patient outcome. **Supplementary Figure 1** show spaghetti plots for both sPLA_2_-V and sPLA_2_-X, which follow a similar pattern of PLA_2_-IIA. A decision tree generated by recursive partitioning assessed whether changes in PLA_2_-IIA levels between day 0 and day 3 (Δ1) and between day 3 and day 7 (Δ2) could stratify patients into mild, severe, and deceased categories (**Figure 3B**). The decision tree corroborated the sPLA_2_-IIA kinetics by first separating the death and severe categories from the mild category using Δ1 values. Patients with a moderate increase in PLA_2_-IIA from day 0 to day 3 (Δ1 ≥ 0.094) were at a significantly higher risk of death and intubation.

**Figure 2:**
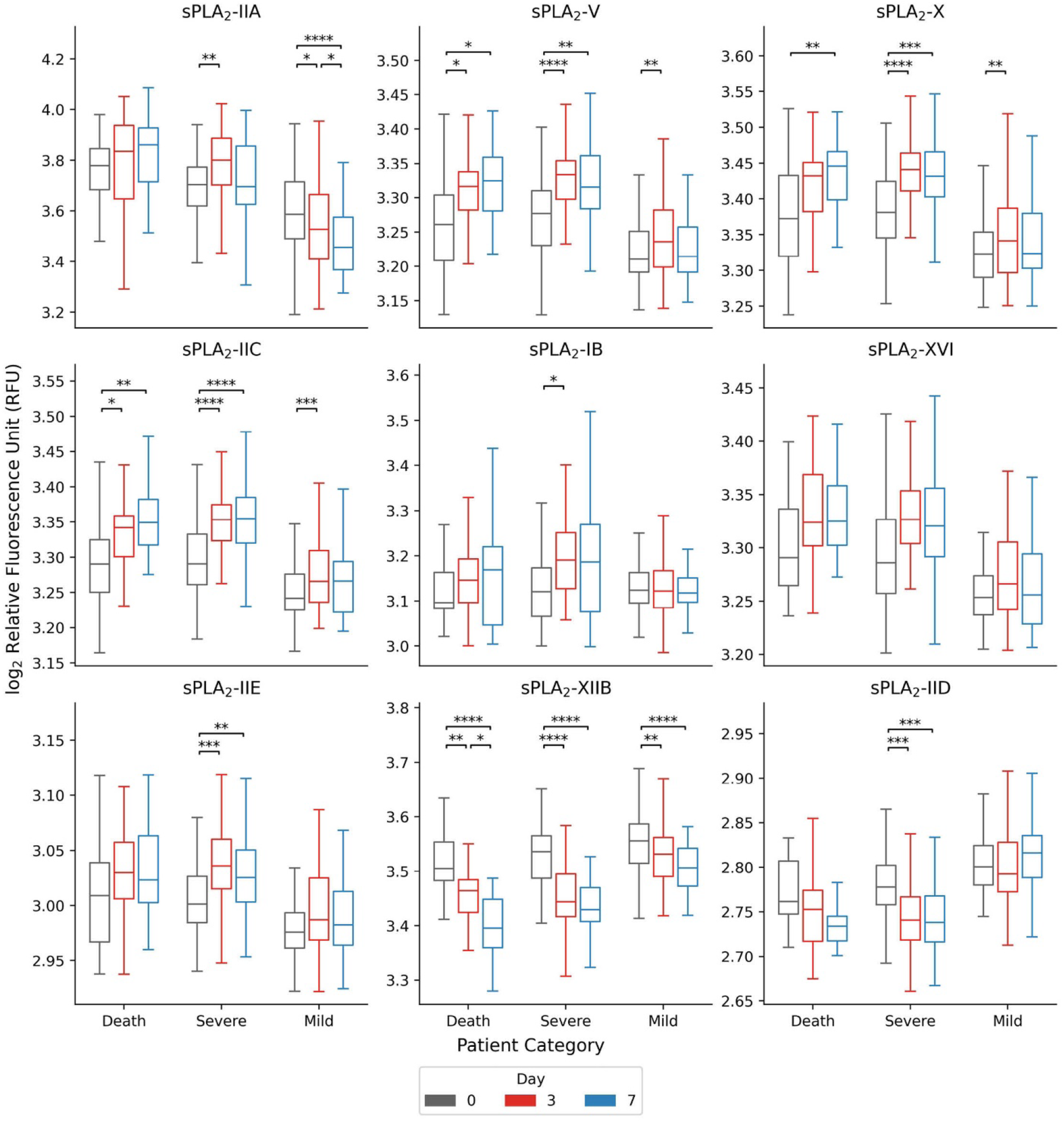
Kinetics of Levels of sPLA_2_ Isoforms in Different Severity Groups. Log_2_ Relative fluorescence units (RFUs) of sPLA_2_ isoform levels by severity over time. Asterisks denote the significance of differences between two days’ values using two-sided independent t-tests. P-values are Bonferroni-adjusted over the three tests conducted within each category. Asterisk p-value ranges provided in methods.

**Figure 3:**
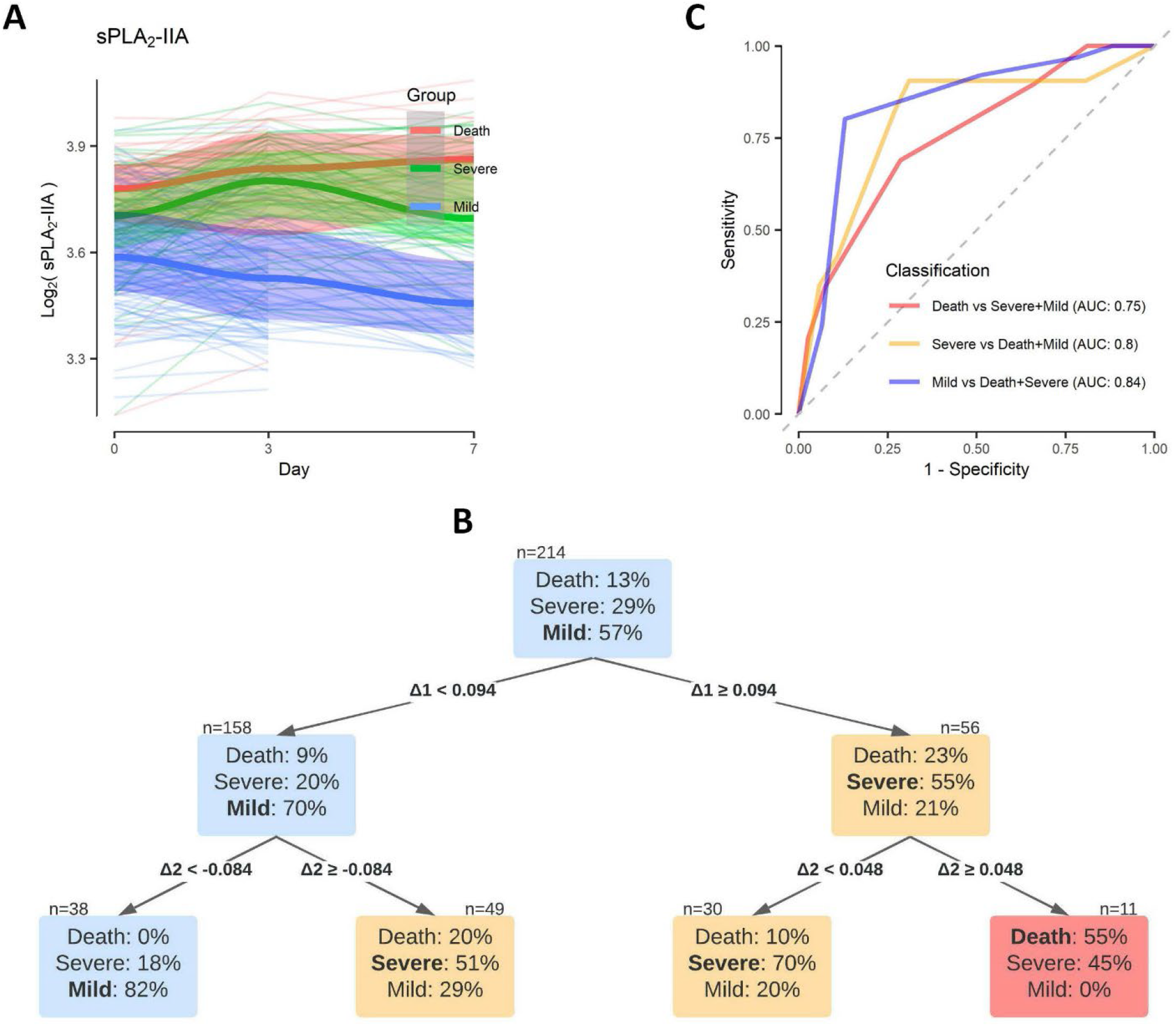
Category Classification Using Changes in sPLA_2_-IIA Levels. Spaghetti plot for longitudinal log_2_ protein levels of sPLA_2_-IIA isoform separated by death, severe, and mild categories for 128 patients with all 3 timepoints. Individuals are plotted as lighter lines and medians as thicker lines. Shaded areas represent the standard deviations of each category. Decision tree stratifying patients from the death, severe, and mild categories. Split decisions are calculated by Gini impurity using day 3 – day 0 and day 7 – day 3 log_2_ relative fluorescence units (RFUs), or Δ1 and Δ 2 values, respectively. Decision values leading to split nodes are labeled on each arrow. The number of samples following a split is labeled above each node. The percentage of each category within the node’s samples are displayed on the node’s face. Node colors are determined by the category with the largest number of samples within the node. **C**. ROC curves and corresponding AUC values reflect the performance of decision trees generated to classify each category alone against the combination of the others (binary classification).

Furthermore, a continuous increase in PLA_2_-IIA from day 3 to day 7 (Δ2 ≥ 0.048) further enriched for death versus severe but survived (right-most branch). In comparison, a further decrease in PLA_2_-IIA from day 3 to day 7 (Δ2 < 0.048) enriched for the mild patient category as depicted on the left-most branch. Tree performance was assessed by classifying each single acuity group (death, severe, or mild) against the combination of the other groups. The corresponding AUCs (area under the ROC curve = 0.75, 0.8, 0.84) suggest that the kinetics of PLA_2_-IIA alone may distinguish between death, severe, and mild disease categories (**Figure 3C**).

**Supplementary Figure 2** compares the kinetics of sPLA_2_-IIA with several plasma cytokines and chemokines. These cytokines and chemokines had the highest-ranking baseline fold changes between the intubated patients + deceased patients versus the milder patient categories seen in **Supplementary Table 1**. In contrast to sPLA_2_-IIA, IL-6, IL-18, and IL-12B median levels were decreased between day 3 and day 7 in deceased patients while CCL22 kinetics show an unclear trend with time and patient severity. A time-dependent increase in the chemokine CCL27 occurred in deceased patients with CCL27 leveling in both intubated and mild categories, suggesting a similar trend to sPLA_2_-IIA. **Supplementary Table 2** lists recursive partitioning classifiers using Δ1 and Δ2 values of the selected cytokines, chemokines, and sPLA_2_ isoforms, all of which performed worse than sPLA_2_-IIA using the same optimization procedure to identify the minimum 10-fold cross-validation error rate.

## Discussion

This study design, unlike our previous one (14), examined sPLA_2_ kinetics and expanded the possible contributions of sPLA_2_ isoforms to COVID-19 disease pathobiology and prognosis. We demonstrate the plasma levels of several sPLA_2_ isoforms, including sPLA_2_-IIA, sPLA_2_-X, sPLA_2_-V, sPLA_2_-IB, and sPLA_2_-XVI, taper in severe and surviving patients, whereas levels continued to increase in non-survivors. A machine learning analysis (decision tree) confirmed that the kinetic behavior of PLA_2_-IIA (Δ1 and Δ2 values) can separate patients into their respective acuity categories. Importantly, this study also suggests that other isoforms sPLA_2_-V, sPLA_2_-X, sPLA_2-_IB, and sPLA_2_-XVI may underlie lethal COVID-19 disease mechanisms (**Figure 4**). sPLA_2_-V expression in the lung is elevated in asthma patients and asthma mice models, where it localizes to epithelial cells, mast cells, and alveolar macrophages (20-23). Interestingly, *Pla2g5 -/-*knockout mice are protected from alveolar injury after antigen, LPS, or ventilator challenge, suggesting a key role in lung inflammation (24, 25). sPLA_2_-V also may promote antigen presentation to T cells and hydrolyze pulmonary surfactant (26-28).

**Figure 4:**
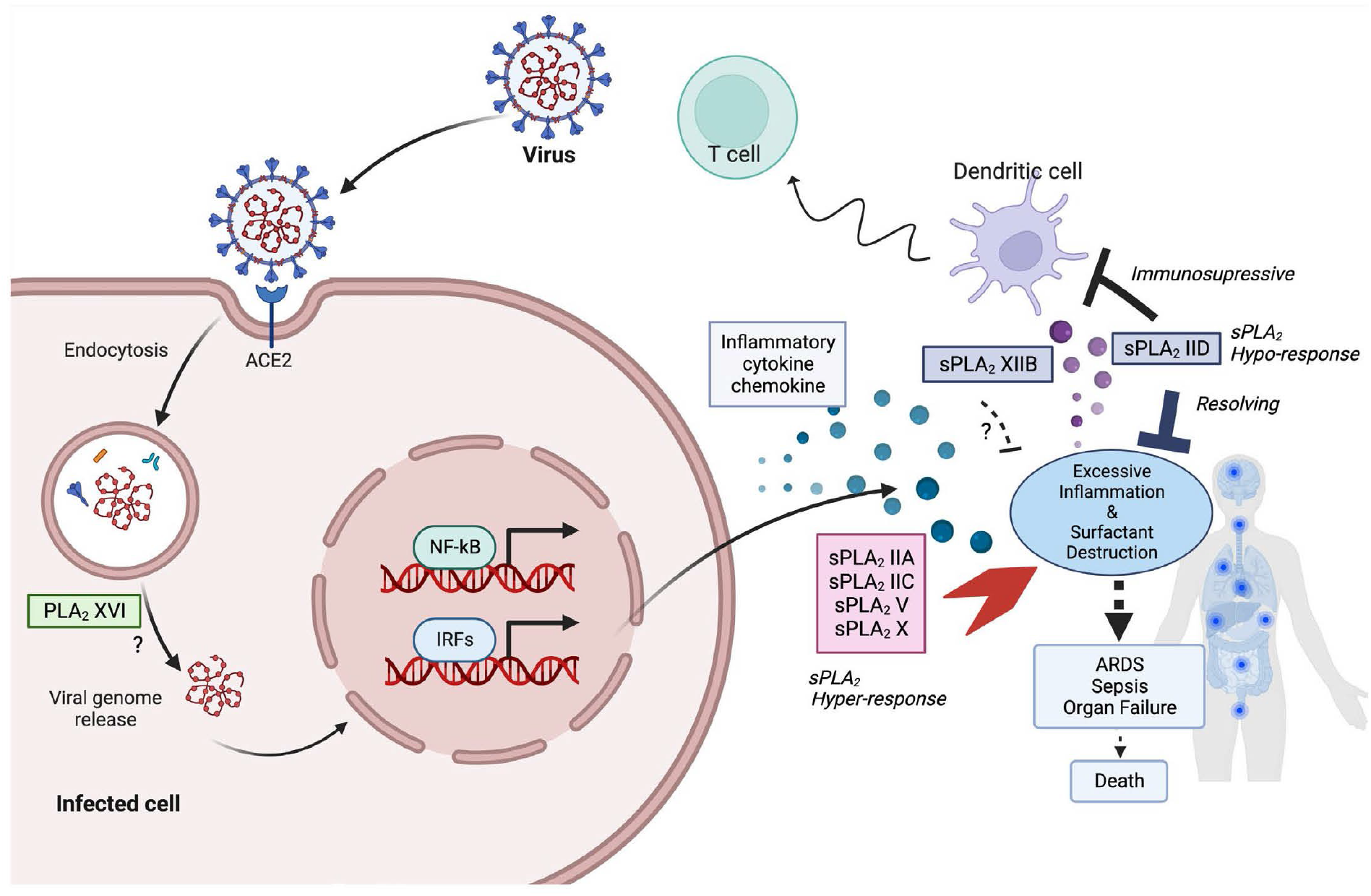
Possible Involvement of the sPLA_2_ Isoforms in COVID-19. The current study shows a persistent increase in sPLA_2_-IIA, -IIC, -V, and -X in deceased patients and lower levels in milder categories. In contrast, the levels of sPLA_2_-IID and -XIIB decrease with lethality and are elevated in milder categories. Based on current knowledge of the function of each isoform, elevated sPLA_2_s are known to have pro-inflammatory effects and thus we are labeling the persistent increase in these as a hyper-responsive sPLA_2_ as their non-regulated increase would likely lead to excessive inflammation, leading to systemic organ failure. In contrast, sPLA_2_-IID has been suggested to have immunosuppressive effects as well as inducing omega-3 fatty acid hydrolysis and subsequent metabolism, thereby resolving inflammation. Thus, we propose the existence of a hypo-response with sPLA_2_-IID as it is reduced in deceased patients, suggesting that its attenuation plays a role in the severity of COVID-19. Little is known about the function of sPLA_2_-XIIB, but its kinetics imply that it may also play an important role in reducing inflammation. sPLA_2_-XVI was previously reported to be a requirement for picornaviruses to release their viral genome into the cytoplasm during infection. Despite picornaviruses being non-enveloped viruses, the kinetics of sPLA_2_-XVI suggest that it could play a role in SARS-CoV-2 infectivity.

Because of sPLA_2_-X’s affinity for phosphatidylcholine and arachidonic acid, much of its proinflammatory properties has been attributed to arachidonic acid derived mediators. sPLA_2_-X occurs in airway epithelial and mast cells and it hydrolyzes pulmonary surfactant (22). *Pla2g10*^−/−^ mice are protected against antigen-induced asthma showing marked reductions in eosinophils, smooth muscle layer thickening, and eicosanoid biosynthesis (20, 22, 29, 30), and these asthmatic responses are restored by a knock-in of the human *PLA2G10* gene (30). sPLA_2_-X has pro-tumorigenic activity in B cell lymphoma via its capacity to hydrolyze highly unsaturated fatty acids such as docosahexaenoic acid from extracellular vesicles; these fatty acids in turn are converted to oxylipin metabolites that promote tumor growth (31).

This present study also identified time dependent reductions in two sPLA_2_ isoforms, sPLA_2_-IID and sPLA_2_-XIIB, in severe COVID-19 disease, with overall higher levels in milder patients. This observation is interesting given that sPLA_2_-IID is a ‘resolving sPLA_2_ isoform’ (**Figure 4**) that mobilizes omega-3 highly unsaturated fatty acids, such as docosahexaenoic acid, that then can serve as precursors for anti-inflammatory/pro-resolving metabolites, such as resolvin D_1_ (32-36). There is an age-dependent increase in this isoform that may worsen outcomes in mice infected with the SARS-COV-2 virus (34). Middle-aged knockout mice lacking expression of *PLA2G2D* have less severe disease and death from SARS-COV-2 (37). sPLA_2_-IID is constitutively expressed in dendritic cells and represses immune responses in Th1, Th2 and Th17 suppressing anti-viral and anti-tumor activities (36). Thus, the progressive reduction in levels observed in deceased patients and the increase in milder patients observed here align with animal studies suggesting an important resolving role in human SARS-COV-2 infection.

Little is known about secreted sPLA_2_-XIIB except that it is catalytically inactive due to altered phospholipid binding properties, may regulate HNF4alpha-induced infectivity of hepatitis C, and is down-regulated in cancer (38-42). We find that sPLA_2_-XIIB kinetics mirror that of sPLA_2_-IID and this suggests it may also have an immunosuppressive role in COVID-19 disease (**Figure 4**). PLA_2_-XVI is not a member of the sPLA_2_family and, as a low molecular weight intracellular protein, has phospholipase A_1_, PLA_2,_ and acyl transferase activities. Importantly, this low molecular weight PLA_2_ is thought to play a role in enabling *Picornaviridae* infections by facilitating virion-mediated transfer into the cytoplasm (17). The higher levels and time-dependent increase of this enzyme in deceased and severe patients suggests a role in SARS-COV-2 infectivity.

Limitations of this study include the inability to collect a validation cohort. Blood collections at the later time points skewed toward sicker patients more likely to remain hospitalized. Patients who died before the 7-day collection time point limited the numbers of patients in this group. However even with these limitations, this study defines the kinetics of appearance of nine secreted PLA_2_ isoforms in circulation and suggests that selective sPLA_2_isoform inhibitors could benefit precision medicine by targeting specific sPLA_2_ isoforms, leading to improved COVID-19 systemic disease and mortality.

## Data Availability

All data produced are available online at Data Mendeley.

https://data.mendeley.com/datasets/nf853r8xsj/2

## 5 Data Availability Statement

The datasets analyzed/generated for this study can be found at Data Mendeley and in the supplemental materials, respectively. Additional requests can be made to the corresponding author.

## 7 Author Contributions

FHC conceived and designed the study. EL, AH, SS, JMS, BH, HZ and GY did the statistical analysis and modeling. FHC wrote the initial manuscript draft. EL, CEM, MCS, JCW, HZ, and GY contributed to revisions and discussion.

