## Supplementary figures and images for "Temporal Associations of Plasma Levels of the Secreted Phospholipase A_2_ Family and Mortality in Severe COVID-19"

### Supplementary Figure 1

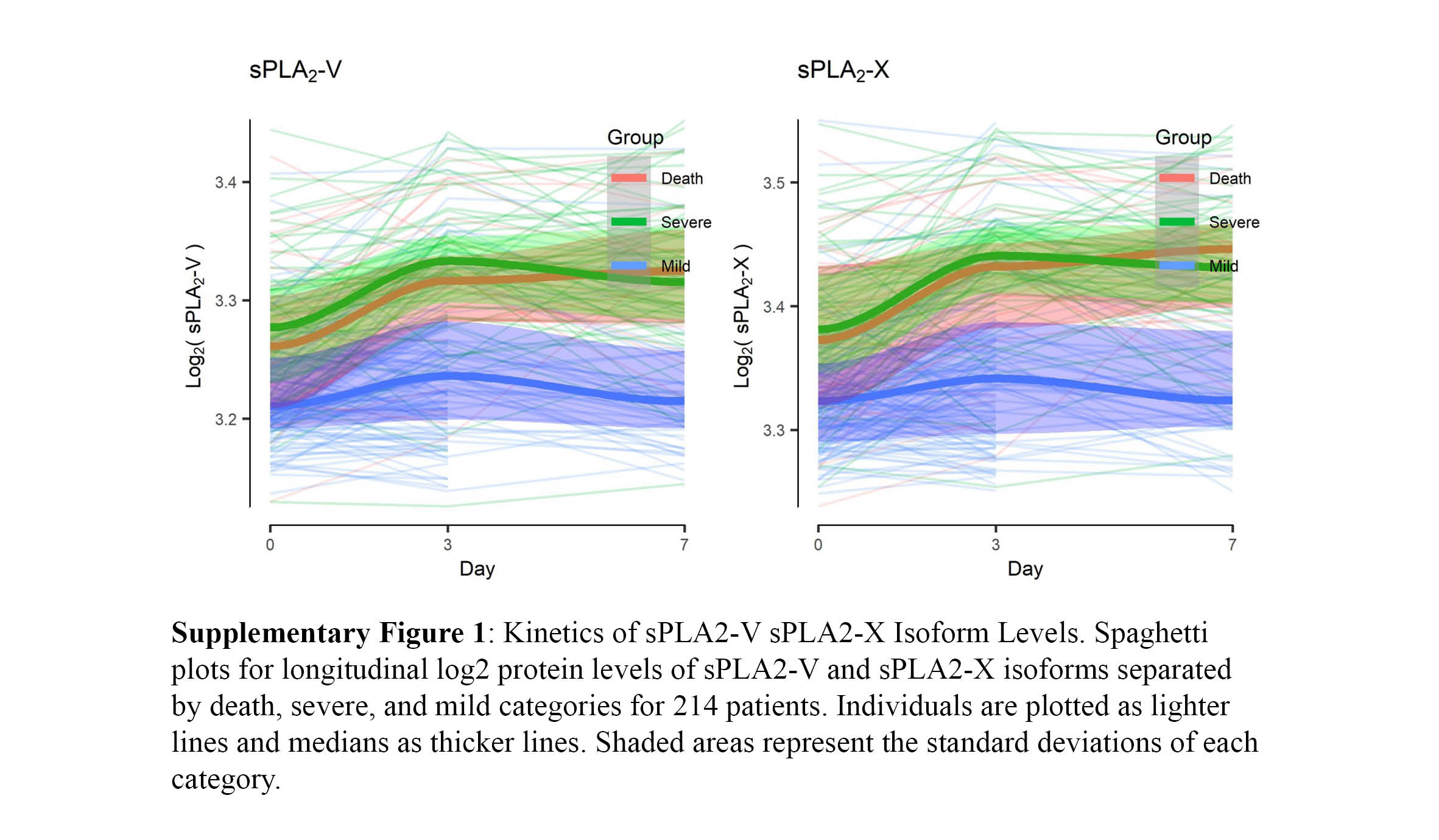

### Supplementary Figure 2

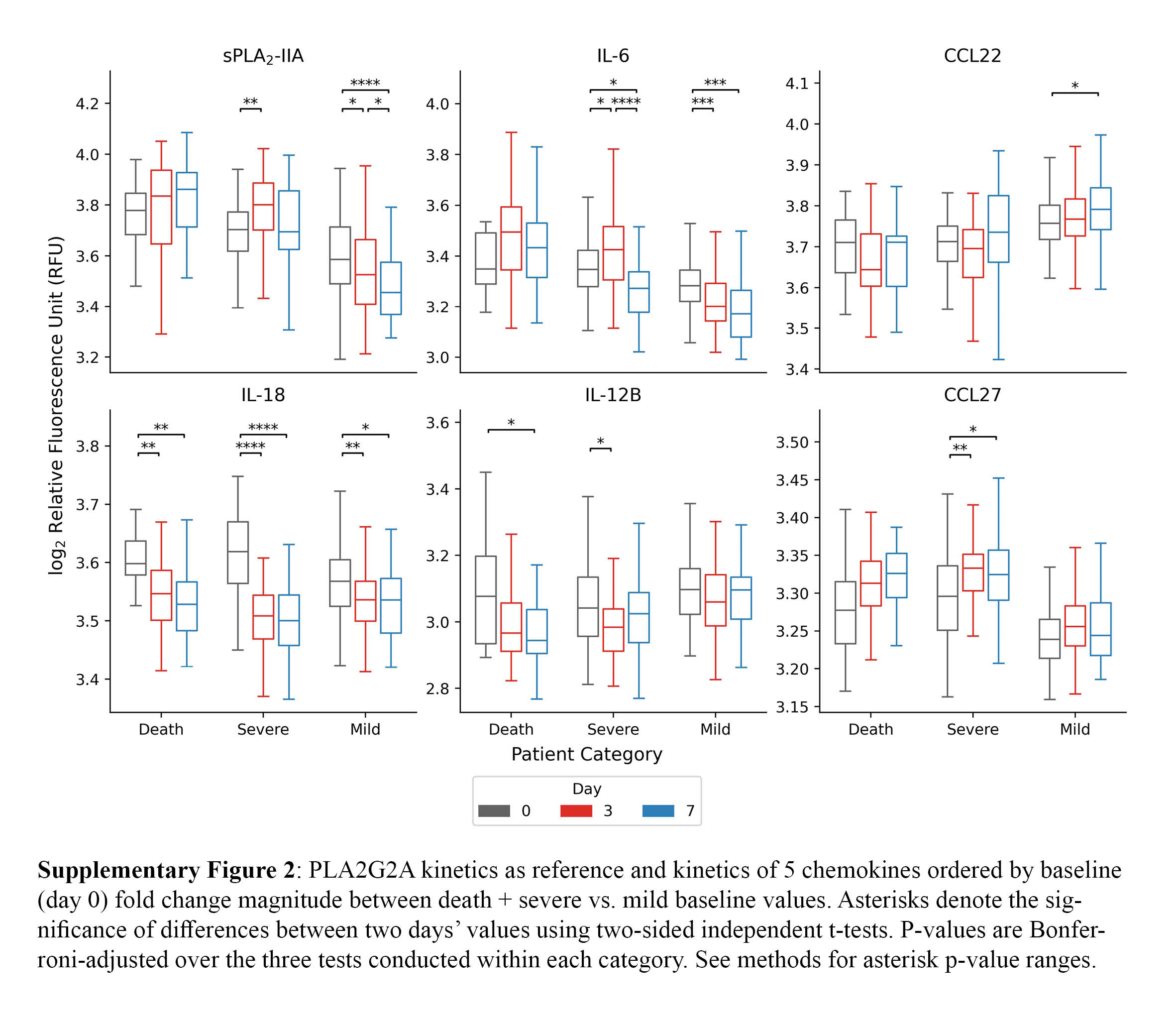
