## Supplementary Table 1 for "Temporal Associations of Plasma Levels of the Secreted Phospholipase A_2_ Family and Mortality in Severe COVID-19"

| Protein | Minimum Cross Validation Error |
| --- | --- |
| sPLA_2_-IIA | 0.30 |
| IL-6 | 0.35 |
| CCL22 | 0.425 |
| IL-18 | 0.39 |
| IL-12B | 0.425 |
| CCL27 | 0.425 |
| sPLA_2_-V | 0.425 |
| sPLA_2_-X | 0.425 |
| sPLA_2_-IIC | 0.425 |
| sPLA_2_-IB | 0.425 |
| sPLA_2_-IIE | 0.425 |
| sPLA_2_-XVI | 0.425 |
| sPLA_2_-XIIB | 0.31 |
| sPLA_2_-IID | 0.37 |

**Supplementary Table 2**: Performance of recursive partition models trained on Δ1 and Δ2 values of specified protein levels reported as 10-fold cross-validation. Under the framework described in the methods, sPLA_2_-IIA outperforms all other sPLA_2_ isoforms and cytokines/chemokines.
